# Convergent structural brain alterations in chronic pain: A multi-metric individual participant data meta-analysis

**DOI:** 10.1101/2025.09.04.25335117

**Authors:** Ryan W.J. Loke, Oscar Ortiz-Angulo, Sylvia M. Gustin, Michèle Hubli, Clas Linnman, Abigail Livny-Ezer, Yann Quidé, Paulina S. Scheuren, John L.K. Kramer

**Affiliations:** Department of Anesthesiology, Pharmacology, and Therapeutics, Faculty of Medicine, University of British Columbia, Vancouver, BC, Canada; School of Biomedical Engineering, Faculty of Applied Sciences, University of British Columbia, Vancouver, BC, Canada; International Collaborations on Repair Discoveries (ICORD), University of British Columbia, Vancouver, BC, Canada; NeuroRecovery Research Hub, School of Psychology, The University of New South Wales (UNSW) Sydney, Sydney, NSW, Australia; Centre for Pain IMPACT, Neuroscience Research Australia, Randwick, NSW, Australia; Spinal Cord Injury Center, Balgrist University Hospital, University of Zurich, Zurich, Switzerland; Neuroscience Center Zurich, ETH Zurich and University of Zurich, Zurich Switzerland; Department of Psychiatry, Massachusetts General Brigham & Harvard Medical School, Boston, USA; Clinical Brain Imaging R&D Center, Sheba Medical Center; Sagol School of Neuroscience, Faculty of Medical Healthy Sciences, Tel Aviv University, Israel

## Abstract

Meta-analyses are a valuable tool in evidence-based research and have contributed to our understanding of structural brain changes in chronic pain. Meta-analyses are also limited by published summary statistics and quality/completeness of underlying studies. To address these limitations, we conducted the first individual participant data (IPD) meta-analysis of brain structure alterations in chronic pain. Using traditional morphometric measures (i.e., volume, cortical thickness, and surface area) and differential-geometric shape metrics (i.e., intrinsic and extrinsic curvature), we aimed to reveal alterations in brain structure convergent across chronic pain conditions. Eight publicly available datasets spanning five conditions and 401 individuals with chronic pain were analyzed: 1) knee osteoarthritis, 2) chronic low back pain, 3) fibromyalgia, 4) migraine, and 5) primary trigeminal neuralgia. FreeSurfer was used to parcellate T1-weighted anatomical images, and metrics for cortical and subcortical structures were extracted. Meta-analysis of study-level comparisons revealed a range of structural changes in the brain associated with chronic pain. Cortical thinning and volume loss were small and localized to the temporo-occipital regions, including bilateral volumetric reductions in the entorhinal cortex. Increases in intrinsic curvature were widespread, involving 49 out of 68 cortical regions. No significant alterations were detected in subcortical volumes. Intrinsic curvature and subcortical volumetric estimates had higher levels of inter-study heterogeneity compared to other metrics, reflecting potential condition and sample specific variability. Leveraging harmonized processing across a large sample size, our novel IPD meta-analysis highlights both widespread and region-specific structural remodeling of chronic pain-related neuroanatomy.

## Introduction

Chronic pain is a leading contributor to all-cause morbidity and disability.^1^ It encompasses biopsychosocial dimensions including sensory, emotional, and behavioral components, that persistently engage a complex network of brain regions.^2,3^ A comparison of brain structure in individuals with chronic pain to healthy sex- and age-matched controls, has revealed clear alterations in brain anatomy. This has been done on a study-by-study basis, revealing marked differences in cortical structure^4–6^ and subcortical volumes^7,8^, then synthesized in conventional, aggregate meta-analyses.^9,10^ While a valuable tool in evidence-based research, conventional meta-analyses rely exclusively on published summary statistics. This introduces biases related to the completeness of reporting and methodological approach taken by the original investigators.^11,12^ Furthermore, heterogeneity across meta-analytic findings, such as decreases^7,8^ and increases^13^ in regional volumes, may reflect condition-specific changes.

Individual participant data (IPD) meta-analyses are now widely embraced as a “gold standard” for meta-analyses.^14^ By processing raw T1-weighted images with a uniform pipeline, IPD meta-analyses reduce variability attributed to methodological heterogeneity (e.g., FreeSurfer vs FSL) and also support subgroup analyses (e.g., sex).^11^ Crucially, with access to raw data, this method allows for the exploration of diverse morphology metrics (e.g., cortical thickness) and differential-geometric measures (intrinsic/extrinsic curvature) that are rarely available in published summaries. The main challenge facing an IPD meta-analysis is access to raw datasets, which is often hampered by data sharing concerns. Despite the abundance of IPD in public repositories like OpenNeuro,^15^ an IPD meta-analysis examining changes brain structure across chronic pain conditions is currently lacking.

Although peripheral mechanisms and clinical phenotypes vary across chronic pain diagnoses, the subjective burden of persistent pain shows cross-diagnostic similarity.^16^ We therefore adopt a cross-condition IPD strategy, utilizing a single, harmonized segmentation pipeline, and pooling across conditions to increase power and detect convergent neuroanatomical differences. In this study, we analyzed structural MRI data from 8 publicly available data sets across five chronic pain conditions: knee osteoarthritis (OA), fibromyalgia (FM), chronic low back pain (CLBP), primary trigeminal neuralgia (PTN), and migraine. We examined a range of structural brain metrics – grey matter volume, cortical thickness, surface area, and both intrinsic and extrinsic curvature – in cortical regions and subcortical volumes parcellated using FreeSurfer. Taking an IPD meta-analysis approach, we hypothesized that chronic pain is associated with convergent changes in brain structure common across multiple chronic pain conditions in comparison to healthy controls.

## Methods

### 2.1 Search Strategy

We initially performed a search on OpenNeuro^15^ and the Open Science Framework^17^ databases using the search terms “MRI” and “chronic pain”. We further queried the journal *Scientific Data* using the same search terms and examined study folders in *OpenPain Project (OPP)* database. Finally, a manual search of published studies that captured anatomical MRI data in a sample of individuals with chronic pain was performed to determine additional sources of data (e.g., Github, Figshare, etc.). We limited the inclusion of data from studies that additionally captured anatomical data from age- and sex-matched healthy controls and were previously published to ensure data quality and provenance.

#### 2.2.1 MRI preprocessing – Cortical Parcellation

Three-dimensional T1-weighted (3D T1w) anatomical MRI images from each participant were downloaded locally and processed using the recon-all function from FreeSurfer (version 7.4.1; https://surfer.nmr.mgh.harvard.edu/, Harvard University, Boston, MA, USA)^18^. Processing was performed on a Windows 11 x64 system using the Windows Subsystem for Linux (WSL) with Ubuntu version 22.04 using in-house bash scripts. Processing involved automated steps for cortical and subcortical parcellation along with quantification of various parameters. In brief, this involves motion correction, skull stripping, and intensity normalization to improve image quality. The brain image is then inflated into a spherical shape for precise alignment to a common space using the Talairach atlas coordinates. FreeSurfer then uses advanced segmentation algorithms to identify brain regions and cortical structures. More specifically, surface-based morphometry parcellated each brain into 68 cortical regions (34 per hemisphere) based on the Desikan-Killiany Atlas.

For each parcellated region, eight structural metrics were automatically extracted, however, only five were used for this study: surface area (SurfArea), gray matter volume (GrayVol), cortical thickness (ThickAvg), mean curvature (MeanCurv, or extrinsic curvature), and Gaussian curvature (GausCurv, or intrinsic curvature). The remaining metrics were excluded due to limited biological relevance (NumVert, FoldInd, and CurvInd).

#### 2.2.2 Subcortical segmentation

The recon-all function also segments subcortical structures and extracts volumetric values for each region. FreeSurfer segments structures like the thalamus, hippocampus, and amygdala by registering the anatomical image to an atlas that provides probabilistic information about the location and shape of these structures (Aseg atlas). This is described in detail by Fisch et al. (2002).^19^

We analyzed bilateral subcortical grey matter structures which include the thalamus, accumbens, caudate, putamen, pallidum, hippocampus, and amygdala. We additionally reported volumes of reference compartments with broad biological relevance which include the brain stem, ventral diencephalon, corpus callosum (anterior, mid-anterior, central, mid-posterior, posterior), cerebellar cortex, cerebellar white matter, and the ventricular system (lateral, 3^rd^, 4^th^, and inferior lateral). We excluded regions that are primarily technical (white matter/non-white matter hypointensities), vascular or ependymal structures (left/right vessel, choroid plexus), optic chiasm, and aggregate measures (e.g., total cerebrospinal fluid), due to their lack of interpretability as independent regions of interest in our study.

### 2.3 Quality Control

The Enhancing NeuroImaging Genetics Through Meta-Analysis Consortium (ENIGMA, https://enigma.ini.usc.edu/) Cortical Quality Control Protocol (version 2.0)^20^ was used to visually inspect cortical parcellations. Cortical measurements (volume, thickness, surface area, mean curvature, and Gaussian curvature) along with other structural metrics (subcortical volumes, ICV, and average cortical thickness) were standardized 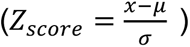 for each region and metric across individuals. Data points with |z| > 3 (∼0.27% tails, conventional threshold for extreme outlier) were flagged for visual inspection and data points with |z| > 5 were auto-excluded as implausible outliers.

### 2.4 Data analysis

Statistical analyses were performed using R version 4.5.1 on a Windows 11 x64 based computer. Each study group was compared to their respective age- and sex-matched healthy controls for each region and parameter. Parametric and non-parametric tests were used according to the results of normality tests (Shapiro-Wilkes test). Effect sizes were calculated using Hedge’s G using the R package esvis (version 0.3.1)^21^ due to the smaller sample sizes of certain studies. The formula Hedge’s g is 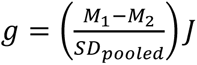, where *M*_1_ – *M*_2_ is the difference in means (i.e. healthy controls – chronic pain). The small sample bias correction factor J is calculated by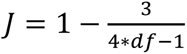 with (*df* = *n*_1_ + *n*_2_ − 2). The standard error for each effect size was also calculated using the following formula, 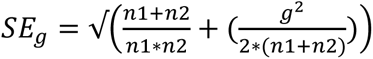 and the upper and lower confidence interval for each effect size was calculated using the following formulas, *CI*_*lower*_ = *Hedges*_*g*_ − 1.96 ∗ *SE*_*g*_ and *CI*_*upper*_ = *Hedges*_*g*_ + 1.96 ∗ *SE*_*g*_. The Benjamini-Hochberg method was used to correct for False Discovery Rate (FDR) with a significance level set to *⍺* = 0.05. P values were adjusted within each metric-specific family (e.g., cortical thickness of all regions or all subcortical volumes).

### 2.5 Meta-Analysis

The meta-analysis was conducted using the metafor package (version 4.8-0).^22^ Using the effect sizes (Hedge’s g) and their corresponding variances previously calculated, a random-effects model was fitted using the rma() function with the restricted maximum likelihood (REML) estimator to account for between-study heterogeneity. The REML method was selected due to its favorable statistical properties, including reduced bias in smaller sample sizes. Heterogeneity was assessed using the *I*^2^ index, *τ*^2^ and Cochrane’s Q and associated p-value for each of the 340 region-metric combinations and 28 subcortical volumes. Cochrane’s Q p-value was also corrected for multiple comparisons using the Benjamini-Hochberg method, with a significance level for adjusted p-values set to *⍺* = 0.05. Prediction intervals were also calculated per region-metric to assess heterogeneity using the r package meta (version 8.1-0).^23^

### 2.6 Sex effects

Datasets were stratified by sex, and analyses were rerun within cohorts from each study. Hedge’s g effect sizes and their respective variances were computed separately for males and females within each study, region, and structural metric. Sex-specific effect size estimates were compared using a two-sided Wald Z test of difference for each region-metric combination. For each male and female effect size estimate per region-metric combination, the formula 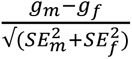 was used to calculate a Wald z-score. Significance was assessed at *⍺* = 0.05 corresponding to |z| > 1.96.

## Results

### 3.1 Dataset breakdown

Eight datasets comprising T1-weighted anatomical MRI scans from seven previously published case-control studies were included in this IPD meta-analysis. These datasets span five chronic pain conditions: knee OA^24^, CLBP^25,26^, FM^27,28^, migraine^29^, and PTN^30^. Each dataset included age- and sex-matched healthy controls. Demographic details and scan acquisition parameters are provided per study in Table 1. Methodological details for each dataset are available in the original publications cited and individual study-level comparative statistics are provided in supplementary tables 1-4.

**Table 1:**
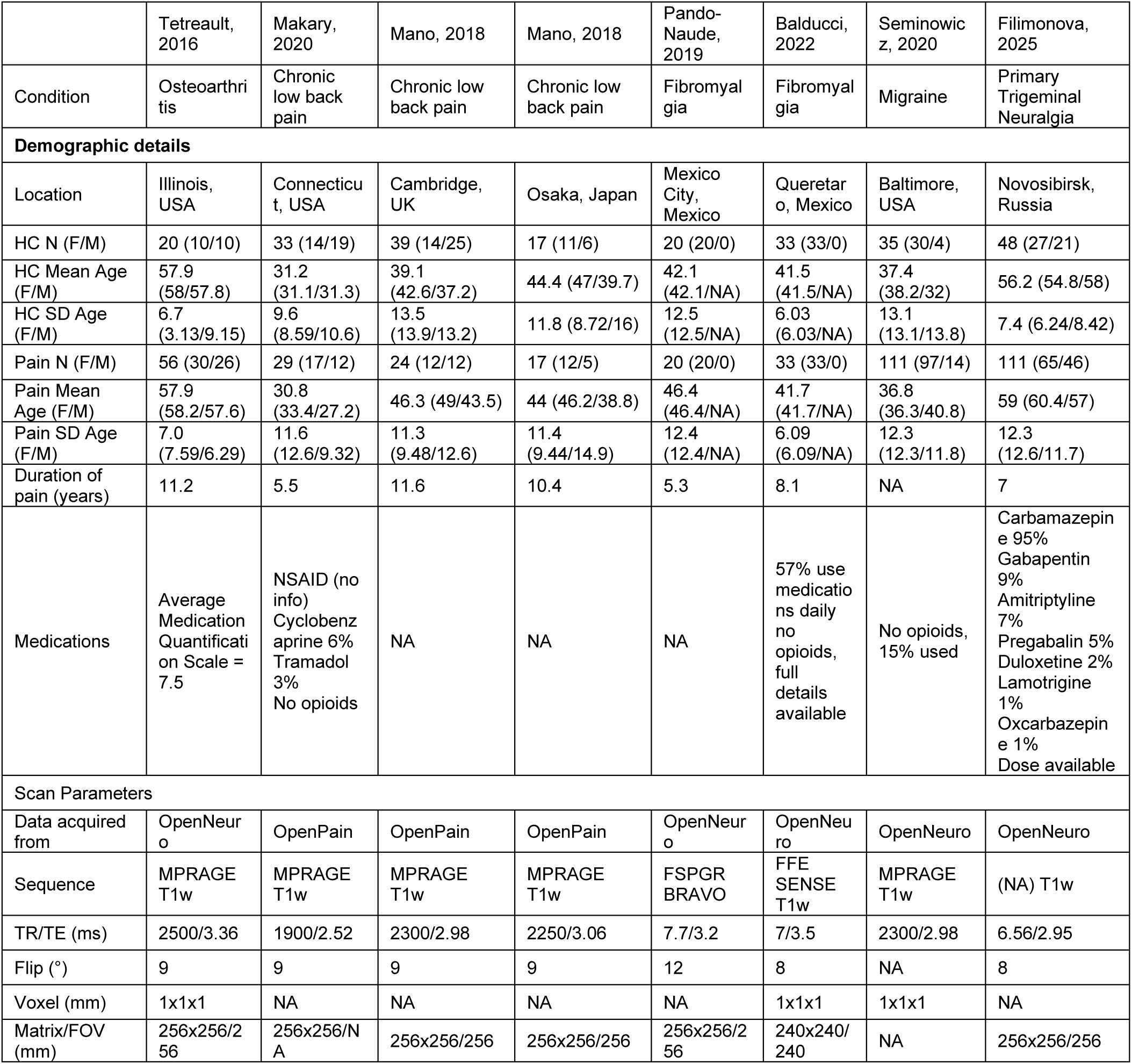
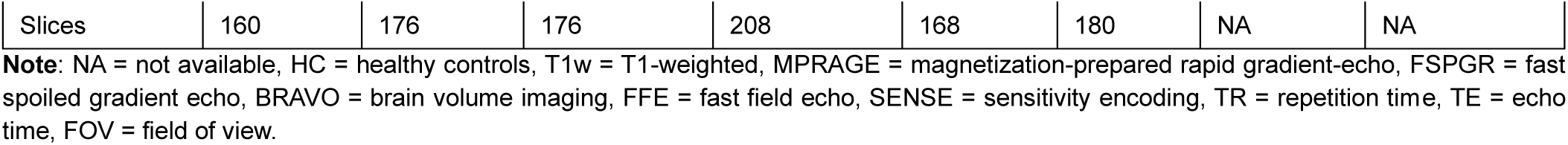
Demographic details and MRI acquisition parameters per data set.

|  | Tetreault, 2016 | Makary, 2020 | Mano, 2018 | Mano, 2018 | Pando-Naude, 2019 | Balducci, 2022 | Seminowicz, 2020 | Filimonova, 2025 |
| --- | --- | --- | --- | --- | --- | --- | --- | --- |
| Condition | Osteoarthritis | Chronic low back pain | Chronic low back pain | Chronic low back pain | Fibromyalgia | Fibromyalgia | Migraine | Primary Trigeminal Neuralgia |
| Demographic details |  |  |  |  |  |  |  |  |
| Location | Illinois, USA | Connecticut, USA | Cambridge, UK | Osaka, Japan | Mexico City, Mexico | Queretaro, Mexico | Baltimore, USA | Novosibirsk, Russia |
| HC N (F/M) | 20 (10/10) | 33 (14/19) | 39 (14/25) | 17 (11/6) | 20 (20/0) | 33 (33/0) | 35 (30/4) | 48 (27/21) |
| HC Mean Age (F/M) | 57.9 (58/57.8) | 31.2 (31.1/31.3) | 39.1 (42.6/37.2) | 44.4 (47/39.7) | 42.1 (42.1/NA) | 41.5 (41.5/NA) | 37.4 (38.2/32) | 56.2 (54.8/58) |
| HC SD Age (F/M) | 6.7 (3.13/9.15) | 9.6 (8.59/10.6) | 13.5 (13.9/13.2) | 11.8 (8.72/16) | 12.5 (12.5/NA) | 6.03 (6.03/NA) | 13.1 (13.1/13.8) | 7.4 (6.24/8.42) |
| Pain N (F/M) | 56 (30/26) | 29 (17/12) | 24 (12/12) | 17 (12/5) | 20 (20/0) | 33 (33/0) | 111 (97/14) | 111 (65/46) |
| Pain Mean Age (F/M) | 57.9 (58.2/57.6) | 30.8 (33.4/27.2) | 46.3 (49/43.5) | 44 (46.2/38.8) | 46.4 (46.4/NA) | 41.7 (41.7/NA) | 36.8 (36.3/40.8) | 59 (60.4/57) |
| Pain SD Age (F/M) | 7.0 (7.59/6.29) | 11.6 (12.6/9.32) | 11.3 (9.48/12.6) | 11.4 (9.44/14.9) | 12.4 (12.4/NA) | 6.09 (6.09/NA) | 12.3 (12.3/11.8) | 12.3 (12.6/11.7) |
| Duration of pain (years) | 11.2 | 5.5 | 11.6 | 10.4 | 5.3 | 8.1 | NA | 7 |
| Medications | Average Medication Quantification Scale = 7.5 | NSAID (no info)<br>Cyclobenzaprine 6%<br>Tramadol 3%<br>No opioids | NA | NA | NA | 57% use medications daily<br>no opioids, full details available | No opioids, 15% used | Carbamazepine 95%<br>Gabapentin 9%<br>Amitriptyline 7%<br>Pregabalin 5%<br>Duloxetine 2%<br>Lamotrigine 1%<br>Oxcarbazepine 1%<br>Dose available |
| Scan Parameters |  |  |  |  |  |  |  |  |
| Data acquired from | OpenNeuro | OpenPain | OpenPain | OpenPain | OpenNeuro | OpenNeuro | OpenNeuro | OpenNeuro |
| Sequence | MPRAGE T1w | MPRAGE T1w | MPRAGE T1w | MPRAGE T1w | FSPGR BRAVO | FFE SENSE T1w | MPRAGE T1w | (NA) T1w |
| TR/TE (ms) | 2500/3.36 | 1900/2.52 | 2300/2.98 | 2250/3.06 | 7.7/3.2 | 7/3.5 | 2300/2.98 | 6.56/2.95 |
| Flip (°) | 9 | 9 | 9 | 9 | 12 | 8 | NA | 8 |
| Voxel (mm) | 1x1x1 | NA | NA | NA | NA | 1x1x1 | 1x1x1 | NA |
| Matrix/FOV (mm) | 256x256/256 | 256x256/NA | 256x256/256 | 256x256/256 | 256x256/256 | 240x240/240 | NA | 256x256/256 |
| Slices | 160 | 176 | 176 | 208 | 168 | 180 | NA | NA |

**Note:** NA = not available, HC = healthy controls, T1w = T1-weighted, MPRAGE = magnetization-prepared rapid gradient-echo, FSPGR = fast spoiled gradient echo, BRAVO = brain volume imaging, FFE = fast field echo, SENSE = sensitivity encoding, TR = repetition time, TE = echo time, FOV = field of view.

### 3.2 IPD Meta-Analysis Results

Significant reductions in grey matter volume and cortical thickness were observed in individuals with chronic pain (Figure 2). Cortical thinning occurred in the right lingual gyrus, right parahippocampal gyrus, right fusiform gyrus, and left transverse temporal gyrus. Volume reductions were detected in the left parahippocampal gyrus, right lingual gyrus, and bilaterally in the entorhinal cortex – notably, the only region to show bilateral alterations across classic structural metrics. There were no significant changes in surface area across the cortex.

**Figure 1:**
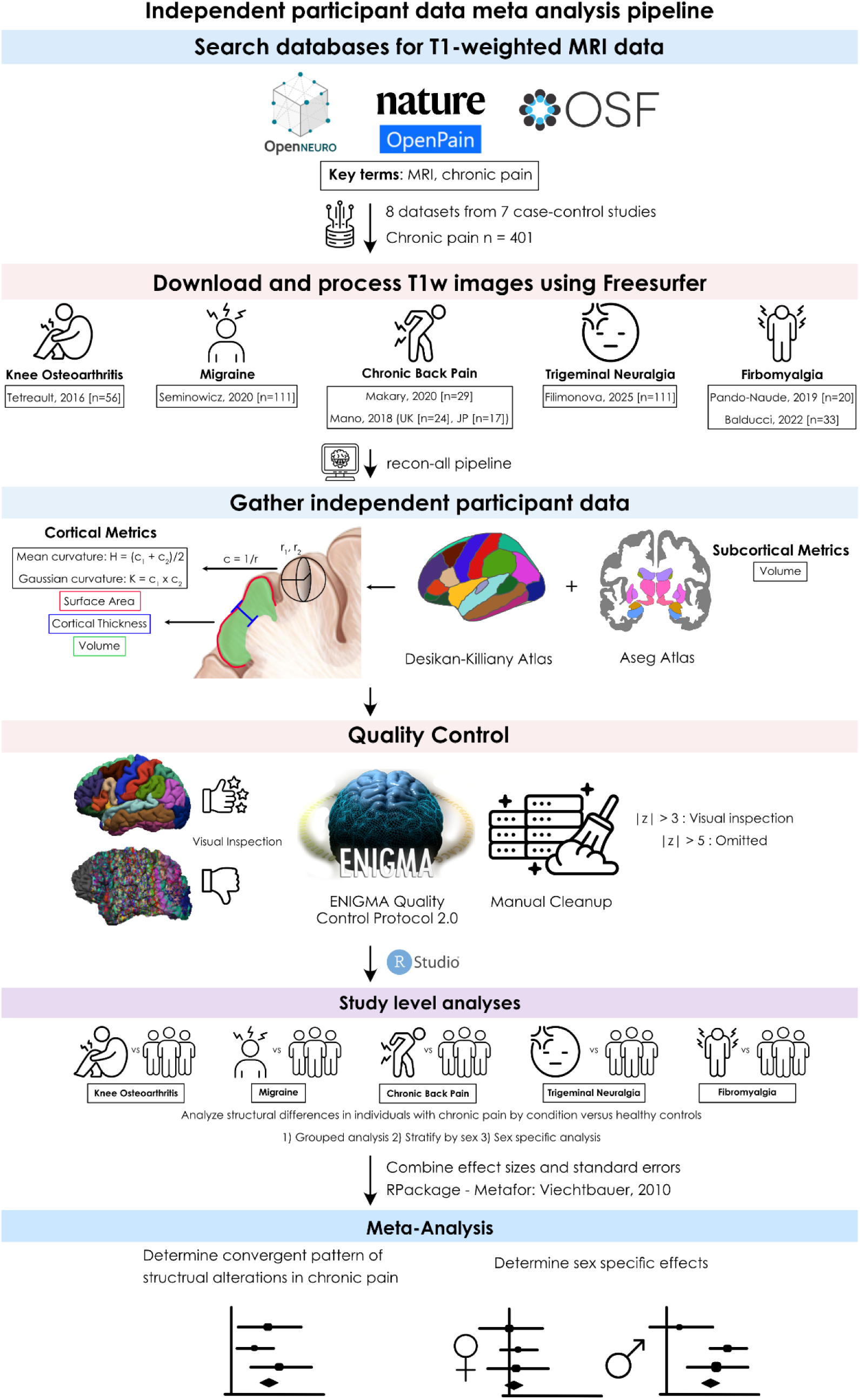
Flow diagram of methodological pipeline. The process includes database searching, processing of images, data extraction, quality control, study level analysis, and meta-analysis.

**Figure 2:**
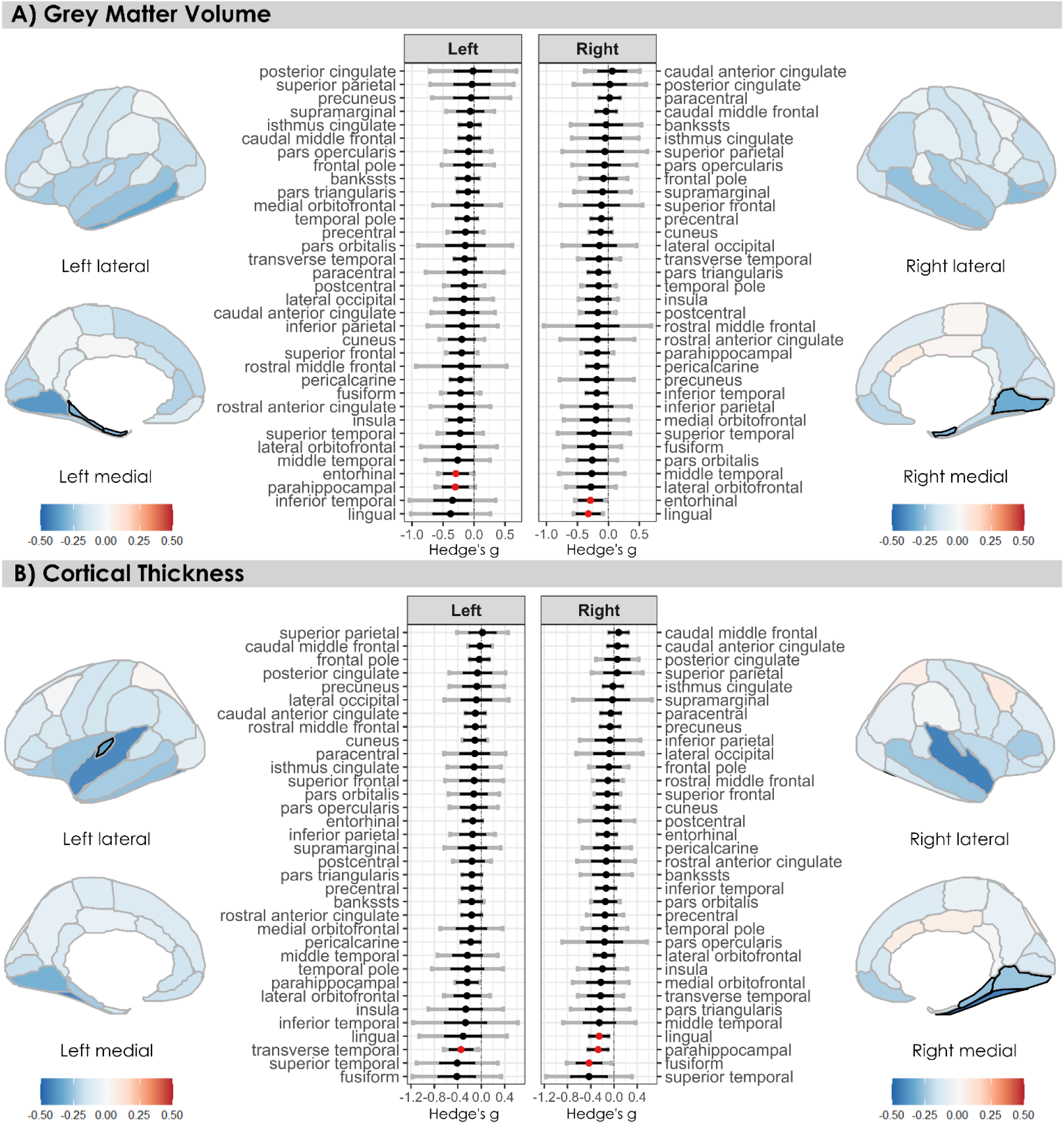
Estimated effect sizes (Hedge’s g) for volume (A) and cortical thickness (B) per region, colored by significance (red dot/black outline = FDR p < 0.05). Positive effect sizes (red) indicate larger values in individuals with chronic pain and negative effect sizes (blue) indicate smaller values in individuals with chronic pain. Black error bars indicate 95% confidence intervals and grey error bars indicate prediction intervals.

We observed significant increases in both intrinsic and extrinsic curvature in individuals with chronic pain (Figure 3). Intrinsic curvature effects were widespread (49 regions, 21 of which were affected bilaterally), whereas extrinsic curvature effects were restricted to the left lateral orbitofrontal cortex, the left temporal pole, and the right caudal anterior cingulate cortex. All meta-analysis statistics are provided in supplementary table 5.

**Figure 3:**
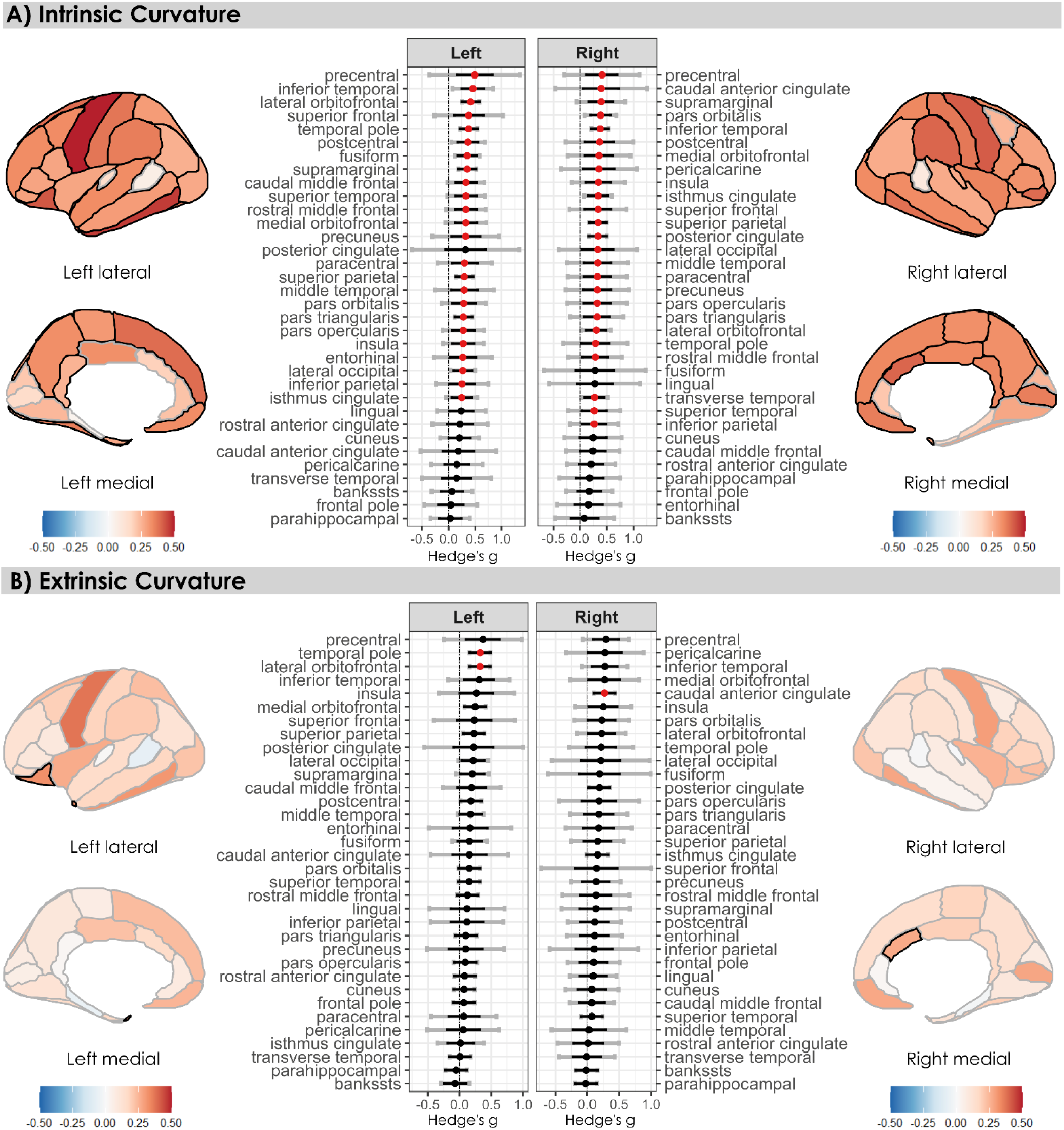
Estimated effect sizes (Hedge’s g) for intrinsic (A) and extrinsic curvature (B) per region, colored by significance (red dot/black outline = FDR p < 0.05,). Positive effect sizes (red) indicate larger values in individuals with chronic pain and negative effect sizes (blue) indicate smaller values in individuals with chronic pain. Black error bars indicate 95% confidence intervals and grey error bars indicate prediction intervals.

Overall, between-study heterogeneity was low to moderate. Significant heterogeneity (Cochrane’s Q, adjusted p < 0.05) was observed in 7 region-metric combinations, 5 of which include the intrinsic curvature of the left posterior cingulate, right fusiform, right caudal anterior cingulate, right lingual cortex, and the left precentral gyrus. The remaining two were the extrinsic curvature of the superior frontal gyrus and the volume of the rostral middle frontal gyrus. Heterogeneity statistics are provided in supplementary table 5.

No significant alterations were detected in subcortical volumes in individuals with chronic pain (Figure 4). Effect size estimates were generally negligible, (|g| < 0.2) except for the amygdala, hippocampus, and cerebellar cortex (0.2 < |g| < 0.4), and were symmetrical across the left and right hemisphere (e.g., the left and right amygdala share similar effect sizes in magnitude and directionality).

**Figure 4:**
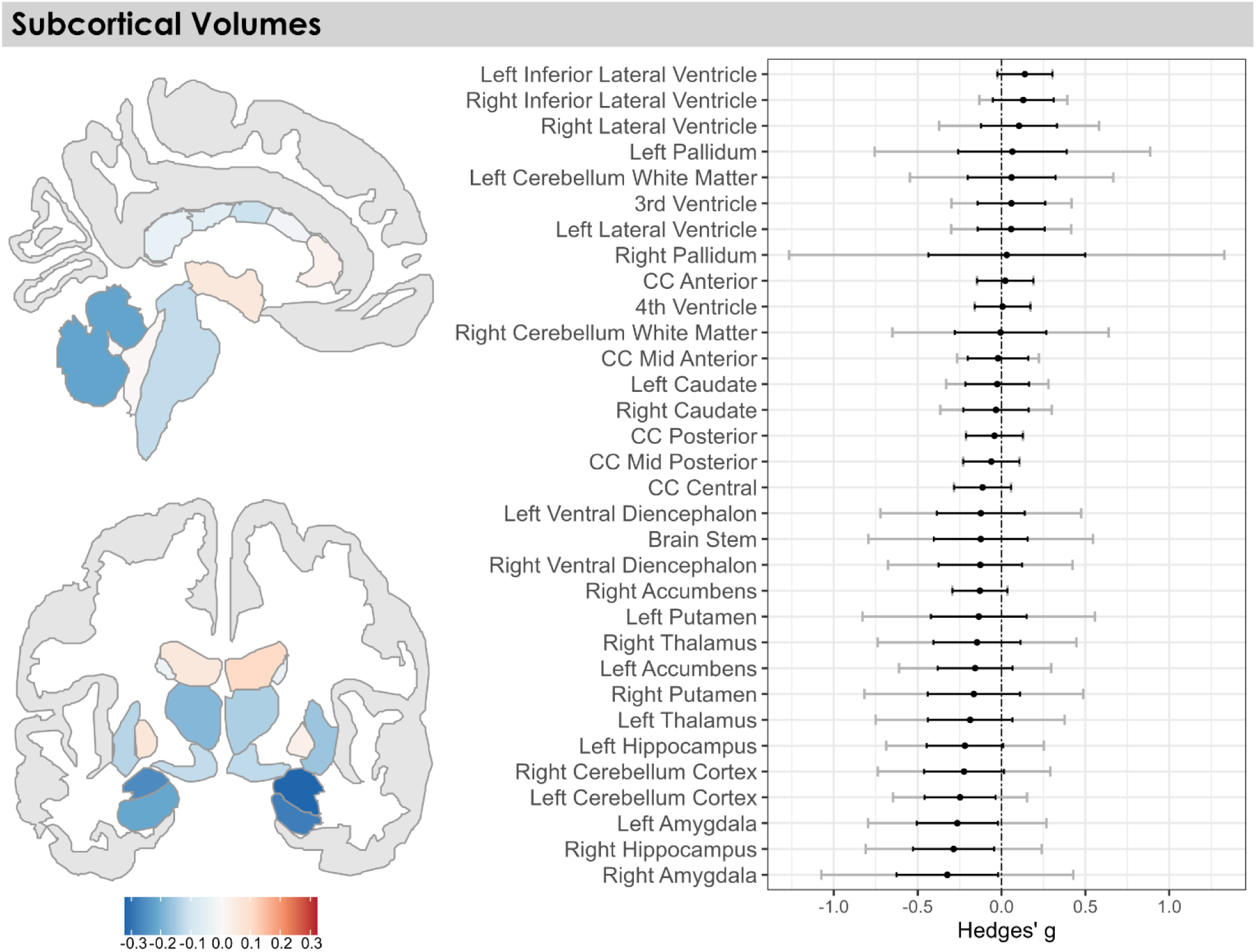
Estimated effect size for each subcortical region, colored for significance following FDR correction (red dot/black outline = p < 0.05, black dot/grey outline = p > 0.05). Positive effect sizes (red) indicate larger volumes in individuals with chronic pain and negative effect sizes (blue) indicate smaller volumes in individuals with chronic pain. Black error bars indicate 95% confidence intervals and grey error bars indicate prediction intervals. Not pictured in the brain plot: the nucleus accumbens, the inferior lateral ventricle, and the ventral diencephalon.

Five subcortical regions showed significant heterogeneity, with the right and left pallidum showing the widest prediction intervals (right: [-1.60, 1.53], left: [-1.06, 0.93]). Other regions displaying significant heterogeneity were the right amygdala, left putamen, and the brain stem. Heterogeneity statistics are provided in supplementary table 5.

### 3.3 Sex effects

Within individual brain regions, there were no significant sex effects for any of the structural or geometric metrics. There were similarly no significant regional sex effects for subcortical volumes. Statistics for sex effects are reported in supplementary table 5.

### 3.4 RShiny App

We developed a publicly accessibly RShiny web application that allows the users to explore the raw data, study-level comparisons, and results from the IPD meta-analysis. (https://lokeryan.shinyapps.io/chronic_pain/)

## Discussion

The aim of our study was to determine changes in brain morphology associated with chronic pain independent of specific etiologies. We applied identical preprocessing pipelines and quality inspection across eight publicly available T1-weighted MRI datasets. We then performed an IPD meta-analysis of surfaced based brain metrics, thereby mitigating analytic heterogeneity, and incomplete reporting biases baked into conventional aggregate data meta-analyses. For cortical thickness, surface area, and volume, our results point to a restricted number of cortical regions that were only minorly affected by chronic pain. In contrast, chronic pain was associated with widespread, nearly global increases in intrinsic curvature. All significant effects exceeded the threshold for a small effect size (|g| > 0.2) but remained below the threshold for a medium effect size (|g| < 0.5). These results provide new insights into the impact of chronic pain on brain structure, highlighting the potential value of brain curvatures as a novel biomarker of disease.

### 4.1 Conventional metrics: volume, surface area, and cortical thickness

The results of our IPD analysis generally aligned with those of previously published aggregate meta-analyses examining conventional volumetric, surface area and thickness metrics. The overall direction (i.e., reductions) and effect magnitude (i.e., small) of chronic pain on brain structure is in close agreement with a recent anatomic likelihood estimate analysis.^10^ In this recent work, significant effects were only reported after relaxing statistical thresholds. Other similarities emerged – for example, both aggregate and our IPD meta-analysis indicate a prominent role for the parahippocampal gyrus – a limbic brain structure - in chronic pain.^31^

Volumetric reductions of the entorhinal cortex in individuals with chronic pain represents a bilateral morphometric change detected by way of IPD meta-analysis, not widely reported elsewhere. The entorhinal cortex plays a central role in episodic memory and serves as a critical hub linking the neocortex and hippocampus.^32^ Its engagement in pain-related learning and memory may underlie its vulnerability as chronic pain has been theorized as a maladaptive memory in which emotional and contextual associations persist beyond tissue healing.^33,34^ This pattern may reflect pathological remodeling driven by persistent activation, metabolic stress, or disrupted connectivity within temporal and limbic circuits; however, causality cannot be inferred from cross-sectional data.

Differences emerged in comparison to aggregate data meta-analyses, however, in that we observed no significant associations between subcortical volumes and chronic pain. Reduced thalamus^7,13,31^ and amygdala volumes,^8^ as well as increased volumes in the caudate^7^ and cerebellum^9^ have been reported elsewhere. Our IPD meta-analysis of subcortical volumes yielded high heterogeneity in regions like the pallidum or amygdala, which may reflect condition-specific effects. Larger, multi-condition datasets, such as those being gathered by the ENIGMA-Chronic Pain group are needed to clarify the role of subcortical structures across and within specific chronic pain conditions.^35^

### 4.2 Intrinsic and extrinsic curvature: novel morphometric biomarkers for chronic pain

While changes in traditional morphometric measures like volume, thickness, and surface area provide valuable insights into brain structure, they represent only one aspect of structural remodeling in the brain. To our knowledge, this study is the first to incorporate curvature-based metrics to identify common structural patterns of change across multiple chronic pain conditions.

Geometric properties of the cortical surface, including intrinsic and extrinsic curvature, have emerged as promising but underutilized markers of cortical reorganization with the potential to capture more subtle changes that may be missed by conventional metrics.^36^ Extrinsic curvature reflects the surface’s shape in a 3-dimensional space (e.g., a crumpled piece of paper can be flattened)^37^. Mean or extrinsic curvature corresponds with gyrification of the cortex, with sulci and gyri giving rise to different estimates.^38^ On the basis of this knowledge, increased extrinsic curvature associated with chronic pain may reflect an increase in the number of sulci ‘wrinkling’ in a given region. Our IPD meta-analysis identified a few significant associations with low inter-study heterogeneity, including the right caudal anterior cingulate – a prominent region of the brain thought to be involved in the emotional and affective dimensions of processing pain.^39^

Intrinsic curvature is a fundamental geometric property of the cortical surface that cannot be altered without distortion (e.g., a ball cannot be flattened without tearing)^37^. Gaussian (intrinsic) curvature captures finer scale spatial variations compared to extrinsic curvature, and at the millimeter-centimeter scale, it is thought to reflect tangential organization of cortical connections, with higher curvature reflecting greater short-range projections.^38^ In chronic pain, increases in intrinsic curvature may reflect enhanced local connectivity and reduced long-range integration. For example, increased intrinsic curvature in regions like the postcentral gyrus and insula indicate reinforcement of somatosensory and salience-related circuits, contributing to sensory amplification, pain hypervigilance, and difficulty disengaging from pain.^40,41^ Such patterns parallel functional connectivity findings of strengthened local network coherence and diminished large-scale integration in chronic pain,^42,43^ suggesting a structural correlate of network segregation. The widespread nature of changes in intrinsic curvature – unbound by regions involved in pain processing – may reflect non-specific change in structure associated with disease.

### 4.3 The role of sex on brain structure in chronic pain

To our knowledge, few studies have comprehensively examined the role of sex on brain morphology in individuals with chronic pain. Among studies that have examined the effect of sex, changes in brain structure are reportedly greater in women compared to men.^44^ These studies are limited by small sample sizes, particularly with regards to the number of male participants (see Table 1 in reference),^44^ thus, statistically underpowered. Based on our IPD meta-analysis, brain structure is mostly similar between men and women, insomuch as no single change in region was sexually dimorphic for any given MRI metric.

## Limitations

The main limitation of our study was that we relied entirely on publicly available T1-weighted MRI datasets, some of which heavily skewed towards female participants. Furthermore, we made no attempts to contact authors of data that was not publicly available. The inclusion of other data would be an asset for our study and any further exploration of an IPD meta-analysis. Further, missing from our study was a complete accounting of pain severity accompanying MRI scans. In addition, we did not control for technical factors such as scanner parameters, which may affect segmentation quality and further contribute to the heterogeneity observed across datasets.

Another limitation was the inability to account for long-term analgesic use. Table 1 provides the available medication use data in the included studies. Many analgesics including ibuprofen^45^ and carbamazepine^46^ may alter brain morphometric and can thus potentially explain some of the observed differences. It is, however, difficult to obtain and account for the full medication history in chronic pain patients, as many different interventions are often attempted and discontinued.

## Data Availability

Beyond the current analyses, we developed an RShiny application (https://lokeryan.shinyapps.io/chronic_pain/) that enables interactive visualization of the raw data, study-level comparisons, and meta-analyses. This tool can allow other researchers to explore our analyses in detail and potentially facilitate new lines of inquiry beyond the scope of the present study.

## Conclusion

We performed an IPD meta-analysis examining structural changes in the brain in individuals with diverse chronic pain conditions. Our goal was to elucidate a structural signature of chronic pain. Our approach revealed widespread cortical alterations, including bilateral volumetric reductions in the entorhinal cortex and global increases in intrinsic curvature. These findings support the view of chronic pain as a disorder of large-scale cortical reorganization, extending beyond sensory regions. Notably, the structural patterns identified in our analysis align with functional imaging reports of strengthened local network connectivity and reduced long-range integration, converging on the concept of chronic pain as a disconnected network disease state of the brain. By applying uniform processing and statistical models across datasets, this study offers a standardized framework that can be built upon in future work. Expanding this approach to larger, more diverse cohorts, including longitudinal datasets, will help delineate condition-specific effects and address causal relationships in brain reorganization.

## Supporting information

Supplementary Table 1 Study level cortical results

Supplementary Table 2 Study level cortical results sex stratified

Supplementary Table 3 Study level subcortical results

Supplementary Table 4 Study level subcortical results sex stratified

Supplementary Table 5 Meta results

Supplementary table dictionary

## Data Availability

All data produced in the present study are available upon reasonable request to the authors

https://lokeryan.shinyapps.io/chronic_pain/

https://openneuro.org/datasets/ds000208/versions/1.0.1

https://openneuro.org/datasets/ds001928/versions/1.1.0

https://openneuro.org/datasets/ds004144/versions/1.0.0

https://openpain.org/index.html

https://openneuro.org/datasets/ds005713/versions/1.0.1

https://openneuro.org/datasets/ds005016/versions/1.1.1

## Acknowledgements

A portion of data collection and sharing for this project was provided by the OpenPain Project (OPP; Principal Investigator: A. Vania Apkarian). OPP funding was provided by the National Institute of Neurological Disorders and Stroke (NINDS) and National Institute of Drug Abuse (NIDA). OPP data are disseminated by the Apkarian Lab, Department of Neuroscience at Northwestern University, Chicago.

## References

1. Mills SEE, Nicolson KP, Smith BH. Chronic pain: a review of its epidemiology and associated factors in population-based studies. Br J Anaesth. 2019;123(2):e273–e283. doi:10.1016/j.bja.2019.03.023

2. Vachon-Presseau E, Centeno MV, Ren W, et al. The Emotional Brain as a Predictor and Amplifier of Chronic Pain. J Dent Res. 2016;95(6):605–612. doi:10.1177/0022034516638027

3. Yang S, Chang MC. Chronic Pain: Structural and Functional Changes in Brain Structures and Associated Negative Affective States. Int J Mol Sci. 2019;20(13). doi:10.3390/ijms20133130

4. Hubbard CS, Khan SA, Keaser ML, Mathur VA, Goyal M, Seminowicz DA. Altered Brain Structure and Function Correlate with Disease Severity and Pain Catastrophizing in Migraine Patients. eNeuro. 2014;1(1):ENEURO.0006-14.2014. doi:10.1523/ENEURO.0006-14.2014

5. Jensen KB, Srinivasan P, Spaeth R, et al. Overlapping Structural and Functional Brain Changes in Patients With Long-Term Exposure to Fibromyalgia Pain. Arthritis Rheum. 2013;65(12):3293–3303. doi:10.1002/art.38170

6. Magon S, Sprenger T, Otti A, Papadopoulou A, Gündel H, Noll-Hussong M. Cortical Thickness Alterations in Chronic Pain Disorder: An Exploratory MRI Study. Psychosom Med. 2018;80(7):592–598. doi:10.1097/PSY.0000000000000605

7. Cauda F, Palermo S, Costa T, et al. Gray matter alterations in chronic pain: A network-oriented meta-analytic approach. Neuroimage Clin. 2014;4:676–686. doi:10.1016/j.nicl.2014.04.007

8. Jia Z, Yu S. Grey matter alterations in migraine: A systematic review and meta-analysis. Neuroimage Clin. 2017;14:130–140. doi:10.1016/j.nicl.2017.01.019

9. Shi H, Yuan C, Dai Z, Ma H, Sheng L. Gray matter abnormalities associated with fibromyalgia: A meta-analysis of voxel-based morphometric studies. Semin Arthritis Rheum. 2016;46(3):330–337. doi:10.1016/j.semarthrit.2016.06.002

10. Henn AT, Larsen B, Frahm L, et al. Structural imaging studies of patients with chronic pain: an anatomical likelihood estimate meta-analysis. Pain. 2023;164(1):e10–e24. doi:10.1097/j.pain.0000000000002681

11. Costa ALF, Lopes SLPC. Highlighting the Benefits and Disadvantages of Individual Participant Data Meta-Analysis in Radiology. Radiol Imaging Cancer. 2024;6(2). doi:10.1148/rycan.240018

12. Zwahlen M, Renehan A, Egger M. Meta-analysis in medical research: Potentials and limitations. Urologic Oncology: Seminars and Original Investigations. 2008;26(3):320–329. doi:10.1016/j.urolonc.2006.12.001

13. Pan PL, Zhong JG, Shang HF, et al. Quantitative meta-analysis of grey matter anomalies in neuropathic pain. European Journal of Pain. 2015;19(9):1224–1231. doi:10.1002/ejp.670

14. Veroniki AA, Seitidis G, Tsivgoulis G, Katsanos AH, Mavridis D. An Introduction to Individual Participant Data Meta-analysis. Neurology. 2023;100(23):1102–1110. doi:10.1212/WNL.0000000000207078

15. Markiewicz CJ, Gorgolewski KJ, Feingold F, et al. The OpenNeuro resource for sharing of neuroscience data. Elife. 2021;10. doi:10.7554/eLife.71774

16. Gustin SM, Wilcox SL, Peck CC, Murray GM, Henderson LA. Similarity of suffering: Equivalence of psychological and psychosocial factors in neuropathic and non-neuropathic orofacial pain patients. Pain. 2011;152(4):825–832. doi:10.1016/j.pain.2010.12.033

17. Foster ED, Deardorff A. Open Science Framework (OSF). Journal of the Medical Library Association. 2017;105(2). doi:10.5195/jmla.2017.88

18. Fischl B. FreeSurfer. Neuroimage. 2012;62(2):774–781. doi:10.1016/j.neuroimage.2012.01.021

19. Fischl B, Salat DH, Busa E, et al. Whole Brain Segmentation. Neuron. 2002;33(3):341–355. doi:10.1016/S0896-6273(02)00569-X

20. Stein JL, Medland SE, Vasquez AA, et al. Identification of common variants associated with human hippocampal and intracranial volumes. Nat Genet. 2012;44(5):552–561. doi:10.1038/ng.2250

21. Anderson D. esvis: Visualization and Estimation of Effect Sizes. CRAN: Contributed Packages. Preprint posted online August 13, 2017. doi:10.32614/CRAN.package.esvis

22. Viechtbauer W. Conducting Meta-Analyses in *R* with the **metafor** Package. J Stat Softw. 2010;36(3). doi:10.18637/jss.v036.i03

23. Schwarzer G. meta: General Package for Meta-Analysis. CRAN: Contributed Packages. Preprint posted online February 8, 2006. doi:10.32614/CRAN.package.meta

24. Tétreault P, Mansour A, Vachon-Presseau E, Schnitzer TJ, Apkarian AV, Baliki MN. Brain Connectivity Predicts Placebo Response across Chronic Pain Clinical Trials. PLoS Biol. 2016;14(10):e1002570. doi:10.1371/journal.pbio.1002570

25. Makary MM, Polosecki P, Cecchi GA, et al. Loss of nucleus accumbens low-frequency fluctuations is a signature of chronic pain. Proc Natl Acad Sci U S A. 2020;117(18):10015–10023. doi:10.1073/pnas.1918682117

26. Mano H, Kotecha G, Leibnitz K, et al. Classification and characterisation of brain network changes in chronic back pain: A multicenter study. Wellcome Open Res. 2018;3:19. doi:10.12688/wellcomeopenres.14069.2

27. Pando-Naude V, Barrios FA, Alcauter S, et al. Functional connectivity of music-induced analgesia in fibromyalgia. Sci Rep. 2019;9(1):15486. doi:10.1038/s41598-019-51990-4

28. Balducci T, Rasgado-Toledo J, Valencia A, van Tol MJ, Aleman A, Garza-Villarreal EA. A behavioral and brain imaging dataset with focus on emotion regulation of women with fibromyalgia. Sci Data. 2022;9(1):581. doi:10.1038/s41597-022-01677-9

29. Seminowicz DA, Burrowes SAB, Kearson A, et al. Enhanced mindfulness-based stress reduction in episodic migraine: a randomized clinical trial with magnetic resonance imaging outcomes. Pain. 2020;161(8):1837–1846. doi:10.1097/j.pain.0000000000001860

30. Filimonova E, Pashkov A, Moysak G, Martirosyan A, Rzaev J. Hippocampal Subfield Abnormalities in Patients With Primary Trigeminal Neuralgia. Journal of Neuroimaging. 2025;35(1). doi:10.1111/jon.70026

31. Smallwood RF, Laird AR, Ramage AE, et al. Structural Brain Anomalies and Chronic Pain: A Quantitative Meta-Analysis of Gray Matter Volume. J Pain. 2013;14(7):663–675. doi:10.1016/j.jpain.2013.03.001

32. Witter M. Entorhinal cortex. Scholarpedia. 2011;6(10):4380. doi:10.4249/scholarpedia.4380

33. Mansour AR, Farmer MA, Baliki MN, Apkarian AV. Chronic pain: The role of learning and brain plasticity. Restor Neurol Neurosci. 2014;32(1):129–139. doi:10.3233/RNN-139003

34. Apkarian AV. Pain perception in relation to emotional learning. Curr Opin Neurobiol. 2008;18(4):464–468. doi:10.1016/j.conb.2008.09.012

35. Quidé Y, Jahanshad N, Andoh J, et al. ENIGMA-Chronic Pain: a worldwide initiative to identify brain correlates of chronic pain. Pain. 2024;165(12):2662–2666. doi:10.1097/j.pain.0000000000003317

36. Deppe M, Marinell J, Krämer J, et al. Increased cortical curvature reflects white matter atrophy in individual patients with early multiple sclerosis. Neuroimage Clin. 2014;6:475–487. doi:10.1016/j.nicl.2014.02.012

37. Demirci N, Holland MA. Cortical thickness systematically varies with curvature and depth in healthy human brains. Hum Brain Mapp. 2022;43(6):2064–2084. doi:10.1002/hbm.25776

38. Ronan L, Pienaar R, Williams G, et al. INTRINSIC CURVATURE: A MARKER OF MILLIMETER-SCALE TANGENTIAL CORTICO-CORTICAL CONNECTIVITY? Int J Neural Syst. 2011;21(05):351–366. doi:10.1142/S0129065711002948

39. Loggia ML, Berna C, Kim J, et al. The Lateral Prefrontal Cortex Mediates the Hyperalgesic Effects of Negative Cognitions in Chronic Pain Patients. J Pain. 2015;16(8):692–699. doi:10.1016/j.jpain.2015.04.003

40. Ke J, Yu Y, Zhang X, et al. Functional Alterations in the Posterior Insula and Cerebellum in Migraine Without Aura: A Resting-State MRI Study. Front Behav Neurosci. 2020;14. doi:10.3389/fnbeh.2020.567588

41. Hong H, Suh C, Namgung E, et al. Aberrant Resting-state Functional Connectivity in Complex Regional Pain Syndrome: A Network-based Statistics Analysis. Exp Neurobiol. 2023;32(2):110–118. doi:10.5607/en23003

42. Mansour A, Baria AT, Tetreault P, et al. Global disruption of degree rank order: a hallmark of chronic pain. Sci Rep. 2016;6(1):34853. doi:10.1038/srep34853

43. Ji Y, Liang X, Pei Y, et al. Disrupted topological organization of brain connectome in patients with chronic low back related leg pain and clinical correlations. Sci Rep. 2025;15(1):7515. doi:10.1038/s41598-025-91570-3

44. Gupta A, Mayer EA, Fling C, et al. Sex-based differences in brain alterations across chronic pain conditions. J Neurosci Res. 2017;95(1-2):604–616. doi:10.1002/jnr.23856

45. Le TT, Kuplicki R, Yeh HW, et al. Effect of Ibuprofen on BrainAGE: A Randomized, Placebo-Controlled, Dose-Response Exploratory Study. Biol Psychiatry Cogn Neurosci Neuroimaging. 2018;3(10):836–843. doi:10.1016/j.bpsc.2018.05.002

46. Parise M, Kubo TTA, Doring TM, Tukamoto G, Vincent M, Gasparetto EL. Cuneus and fusiform cortices thickness is reduced in trigeminal neuralgia. J Headache Pain. 2014;15(1):17. doi:10.1186/1129-2377-15-17

