## Supplementary table dictionary for "Convergent structural brain alterations in chronic pain: A multi-metric individual participant data meta-analysis"

**Supplementary Tables Breakdown**

**Supplementary_1_Study_level_cortical_results**

- 8 total sheets, one for each dataset, with study level comparative statistics for cortical regions for each measurement metric.

**Supplementary_2_Study_level_cortical_results_sex_stratified**

- 12 sheets total sheets, 2 per dataset where cohorts can be split by sex, with study level comparative statistics.

**Supplementary_3_Study_level_subcortical_results**

- 8 total sheets, one for each dataset, with study level comparative statistics for subcortical volumes.

**Supplementary_4_Study_level_subcortical_results_sex_stratified**

- 12 total sheets, 2 per dataset where cohorts can be split by sex, with study level comparative statistics.

**Supplementary_5_Meta_Results**

- 8 total sheets, containing meta-analysis results for grouped and sex stratified cortical/subcortical analyses. Sex effects statistics are included here for cortical and subcortical analyses.

**Supplementary Table Dictionary – Study Level Comparisons (Cortical & Subcortical, Tables 1-4)**

Datasheets

| Datasheet Name | Author/study |
| --- | --- |
| OA | Osteoarthritis: Tetreault, 2016 |
| FM_1 | Fibromyalgia: Pando-Naude, 2019 |
| FM_2 | Fibromyalgia: Balducci, 2022 |
| CLBP_1 | Chronic Low Back Pain: Makary, 2020 |
| CLBP_2_S1 | Chronic Low Back Pain: Mano, 2018 (UK Data) |
| CLBP_3_S2 | Chronic Low Back Pain: Mano, 2018 (Japan Data) |
| Migraine | Migraine: Seminowicz, 2020 |
| PTN | Primary Trigeminal Neuralgia: Filimonova, 2025 |

Sex stratified sheets may end with _M or _F indicating male and female specific statistics per study.

Measurement Types

| Abbreviation | Definition |
| --- | --- |
| SurfArea | Surface Area (mm^2^) |
| GrayVol | Grey Matter Volume (mm^3^) |
| ThickAvg | Average Cortical Thickness (mm) |
| MeanCurv | Mean Curvature – extrinsic curvature (1/mm) |
| GausCurv | Gaussian Curvature – intrinsic curvature (1/mm^2^) |

Column Names

FDR_p – p-value corrected for multiple comparisons, grouped by measurement metric.

Mean_Group1 – Average value per measurement of **HEALTHY CONTROLS**

SD_Group1 – Standard deviation of **healthy control** data

N_Group1 – sample size of healthy controls per dataset

Mean_Group2 – Average value per measurement of **INDIVIDUALS WITH CHRONIC PAIN**

SD_Group2 – Standard deviation of **individuals with chronic pain** data

N_Group2 – Sample size of individuals with chronic pain per dataset

Hedges_g – Effect size (hedge’s g) – positive indicates larger values in individuals with chronic pain and negative indicates values smaller in individuals with chronic pain.

SE_Hedges_g – Standard error of hedge’s g

CI_lower – lower confidence interval of Hedge’s g

CI_upper – upper confidence interval of Hedge's g

**Supplementary Table Dictionary – Meta Analysis (Cortical & Subcortical, Table 5)**

K – number of studies included in the meta-analysis

Estimate – estimated pooled effect size per region and metric

SE – standard error of estimated effect size

Zval - Z statistic for test of pooled effect size

Pval – probability value for significance test of estimated effect size

FDR_p – p-value corrected for multiple comparisons, grouped by measurement metric.

CI_lb – confidence interval, lower bound for estimated effect size

CI_ub – confidence interval, upper bound for estimated effect size

PI_lb – prediction interval, lower bound for estimated effect size

PI_ub - prediction interval, upper bound for estimated effect size

I2 – I^2^, percentage of total variation across studies due to heterogeneity

t2 - $\tau^{2}$ = random-effects variance component for between-study heterogeneity

Q – Cochran’s Q statistic for heterogeneity

pval_Q – p-value for Cochran’s Q test of heterogeneity

FDR_Q_p – Cochran’s Q test p-value corrected for multiple comparisons

Z_diff – z-score calculated from Wald-z test

**NOTE:** For sex analysis sheets, column names will end with an additional “_m” or “_f” indicating e.g. male or female specific estimates.
